# Re-emergence of Yellow Fever Virus in Brazil: Evidence from Forest and peri-urban Settings

**DOI:** 10.1101/2025.04.10.25325467

**Authors:** Valnete das Graças Dantas Andrade, Talita Émile Ribeiro Adelino, Vagner Fonseca, Keldenn Melo Farias Moreno, Luiz Marcelo Ribeiro Tomé, Luiz Augusto Pereira, Debora Glenda Lima de La-Roque, Ana Maria Bispo de Filippis, Daniel Garkauskas Ramos, Dario Brock Ramalho, Kátia Cristina de Lima Furtado, Gleissy Adriane Lima Borges, Livia Caricio Martins, Livia C. V. Frutuoso, Ludmila Oliveira Lamounier, Natália Rocha Guimarães, Patrícia Miriam Sayuri Sato Barros, Priscila Souza de Almeida, Paulo Eduardo de Souza da Silva, Rodrigo Giesbrecht Pinheiro, Rodrigo Guerino Stabeli, Shirley Moreira da Silva Chagas, Sílvia Helena Sousa Pietra Pedroso, Simone Kashima, Solange Gonçalves Penante, Marília Santini de Oliveira, Vinicius Lemes da Silva, Wesley C. Van Voorhis, Edward C. Holmes, José Lourenço, Felipe Campos de Melo Iani, Alberto Simões Jorge Júnior, Marta Giovanetti, Luiz Carlos Junior Alcantara

**Affiliations:** Laboratório Central do Estado, Secretaria de Saúde Pública, Belém, PA, Brazil; Fundação Ezequiel Dias, Belo Horizonte, MG, Brazil; Instituto René Rachou, Fundação Oswaldo Cruz, Belo Horizonte, MG, Brazil; Departamento de Ciências Exatas e da Terra, Universidade do Estado da Bahia, Salvador, BA, Brazil; Universidade Federal de Minas Gerais, Belo Horizonte, MG, Brazil; Laboratório Central de Saúde Pública Dr Giovanni Cysneiros, Goiânia, GO, Brazil; Hemocentro de Ribeirão Preto, Ribeirão Preto, SP, Brazil; Faculdade de Medicina de Ribeirão Preto, Universidade de São Paulo, Ribeirão Preto, SP, Brazil; Laboratório de Arbovírus e Vírus Hemorrágicos, Instituto Oswaldo Cruz, Rio de Janeiro, Rio de Janeiro, Brazil; Coordenacao-Geral de Vigilância de Arboviroses, Brazilian Ministry of Health, Brazil; Força Nacional do SUS, Ministerio da Saude, Brasilia, Federal District, Brazil; Seção de Biologia Molecular I, Laboratório Central do Estado, Secretaria de Saúde Pública, Ministério da Saúde, Belém, PA, Brazil; Evandro Chagas Institute, Para, Brazil; Divisão de Biologia Médica, Laboratório Central do Estado, Secretaria de Saúde Pública, Belém, PA, Brazil; Coordenacao-Geral de Laboratórios de Saúde Pública, Brazilian Ministry of Health, Brazil; Center for Emerging and Re-emerging Infectious Diseases (CERID), Department of Medicine, Division of Allergy & Infectious Diseases, University of Washington, Seattle, WA USA; School of Medical Sciences, University of Sydney, Sydney, NSW, Australia; BioISI (Biosystems and Integrative Sciences Institute), University of Lisbon, Lisbon, Portugal; Universidade Católica Portuguesa, Biomedical Research Center, Lisboa, Portugal; Sciences and Technologies for Sustainable Development and One Health, Università Campus Bio-Medico di Roma, Italy, 00128; Oswaldo Cruz Foundation, Oswaldo Cruz Institute, Rio de Janeiro, Brazil

**Keywords:** Yellow fever virus (YFV), Brazil; Sylvatic transmission, re-emergence, genomic surveillance

## Abstract

Yellow Fever virus (YFV) continues to challenge public health systems across the Americas, despite decades of successful control and the availability of a highly effective vaccine. Following major outbreaks between 2016 and 2022, YFV re-emerged in Brazil in 2023, with a confirmed infection in a non-human primate in São Paulo linked to viral strains circulating in the Midwest region. This reintroduction, combined with increased YFV activity reported across the Americas, signals an ongoing risk of viral persistence and geographic expansion. In this study, we investigate recent YFV dynamics in forest and peri-urban settings, integrating genomic evidence with ecological and spatial data to trace transmission pathways and assess the potential for urban re-emergence. Our findings highlight the need for sustained surveillance in both sylvatic and transitional zones, especially amid fluctuating vaccine coverage and environmental changes that may facilitate spillover events.

## Text

Yellow fever (YF), a mosquito-borne disease caused by Yellow Fever virus (species *Orthoflavivirus flavi*) within the family *Flaviviridae*, continues to pose a significant public health challenge in endemic regions of Africa and the Americas (Lindenbach et al., 2013). The virus persists in nature through two primary transmission cycles: a sylvatic cycle, involving nonhuman primates (NHPs) and forest canopy mosquitoes such as *Sabethes* and *Haemagogus* species, and an urban cycle that has not been documented in Brazil since 1942 (Possas et al., 2018). While eradication is unlikely due to the presence of animal reservoirs, high vaccine coverage remains the most effective approach to avert urban outbreaks. However, despite the availability of a highly effective vaccine, Brazil faced notable YFV re-emergence between 2016 and 2022 (Faria et al., 2018; Giovanetti et al., 2019; Giovanetti et al., 2023). In early 2023, a YFV infection detected in a non-human primate in the state of São Paulo was confirmed through genomic analysis to represent a novel introduction originating from the Midwest region of Brazil, reinforcing concerns regarding continued viral dissemination and transmission risk (Fernandes et al., 2023). Later that year, the Pan-American Health Organization (PAHO) reported a broader rise in YFV activity throughout the Americas, reinforcing concerns over the population persistence of the virus and its potential for wider spread (PAHO, 2025). Herein, we investigated the re-emergence of YFV post-2023, with emphasis on its spread in forest and peri-urban settings.

Our geographic analysis of confirmed human cases between 2023 and 2025 revealed a progressive eastward expansion of YFV circulation (**Figure 1**). In 2023, the majority of cases remained concentrated in western Amazonian regions. However, in 2024 and early 2025, new clusters were identified in central and northern Brazil, with increasing detections in southeastern states, including previously unaffected areas of the states of São Paulo and Minas Gerais. This pattern is consistent with historical observations of YFV dissemination from enzootic Amazonian reservoirs into more populated transitional zones (Giovanetti et al., 2023). The country-level distribution of YFV cases demonstrates a marked increase in transmission across the region, with Colombia experiencing increases in 2024-2025 and Brazil experiencing the sharpest rise in 2025 (**Figure 1b**). At the Brazilian state level, surveillance data highlight substantial increases in both case numbers and YFV-related mortality in Pará and Minas Gerais, which remain areas of concern due to their ecological connectivity and the presence of vulnerable populations with suboptimal vaccine coverage (**Figure 1c**).

**Figure 1.**
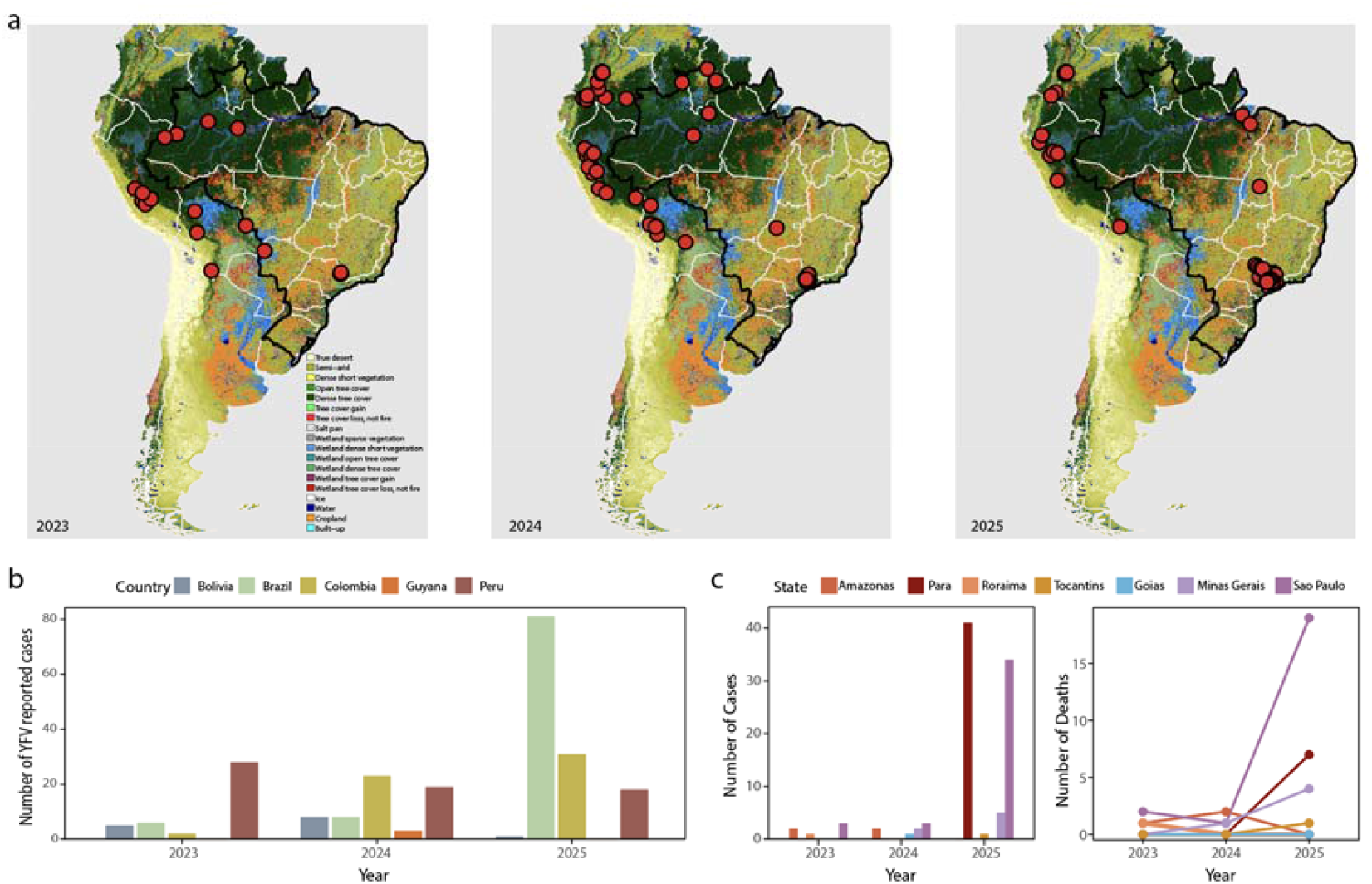
Spatiotemporal dynamics and epidemiological trends of yellow fever virus (YFV) circulation in South America from 2023 to 2025. a) Geographical distribution of confirmed human YFV cases overlaid on a land cover map of South America. Brazil is highlighted with a black border. Red circles indicate reported cases for each respective year (2023, 2024, and 2025); b) Number of confirmed YFV cases reported per year by country (Bolivia, Brazil, Colombia, Guyana, and Peru). A marked increase in total cases is observed in 2025; c) Number of confirmed cases (left) and deaths (right) reported annually by selected Brazilian states. Data highlight rising case numbers in Pará and Minas Gerais, with associated increases in YFV-related mortality.

To investigate the emergence and spread of YFV in two affected Brazilian states - Pará (PA) and Minas Gerais (MG) - we implemented an integrated molecular and genomic surveillance approach. Clinical samples were processed at the state public health laboratories, where viral RNA was extracted and screened by RT-qPCR (Domingo et al., 2012). Positive samples underwent whole-genome sequencing on MiSeq (Illumina) and MinION (Oxford Nanopore) using the COVIDSeq Assay and Native Barcoding Kit 96 V14, respectively. Consensus genomes were aligned with complete YFV sequences from GenBank (as of March 24, 2025) using MAFFT (Katoh and Standley, 2013), and phylogenetic analyses were performed with IQ-TREE2 (Minh et al., 2020) and BEAST v1.10.4 (Brazird et al., 2018).

A total of 29 novel YFV genomes were generated from samples collected between 2023 and 2025, comprising 13 human and 16 non-human primate (NHP) sequences. These genomes were obtained from different sample types, including serum (n = 13), liver (n = 14), spleen (n = 1), and lung (n = 1), and originated from forest, rural, peri-urban, and urban settings (**Table 1**). Samples were geographically distributed between Pará in the Northern region (n = 11) and Minas Gerais in the Southeastern region of Brazil (n = 18) (**Figure 2a**).

**Figure 2.**
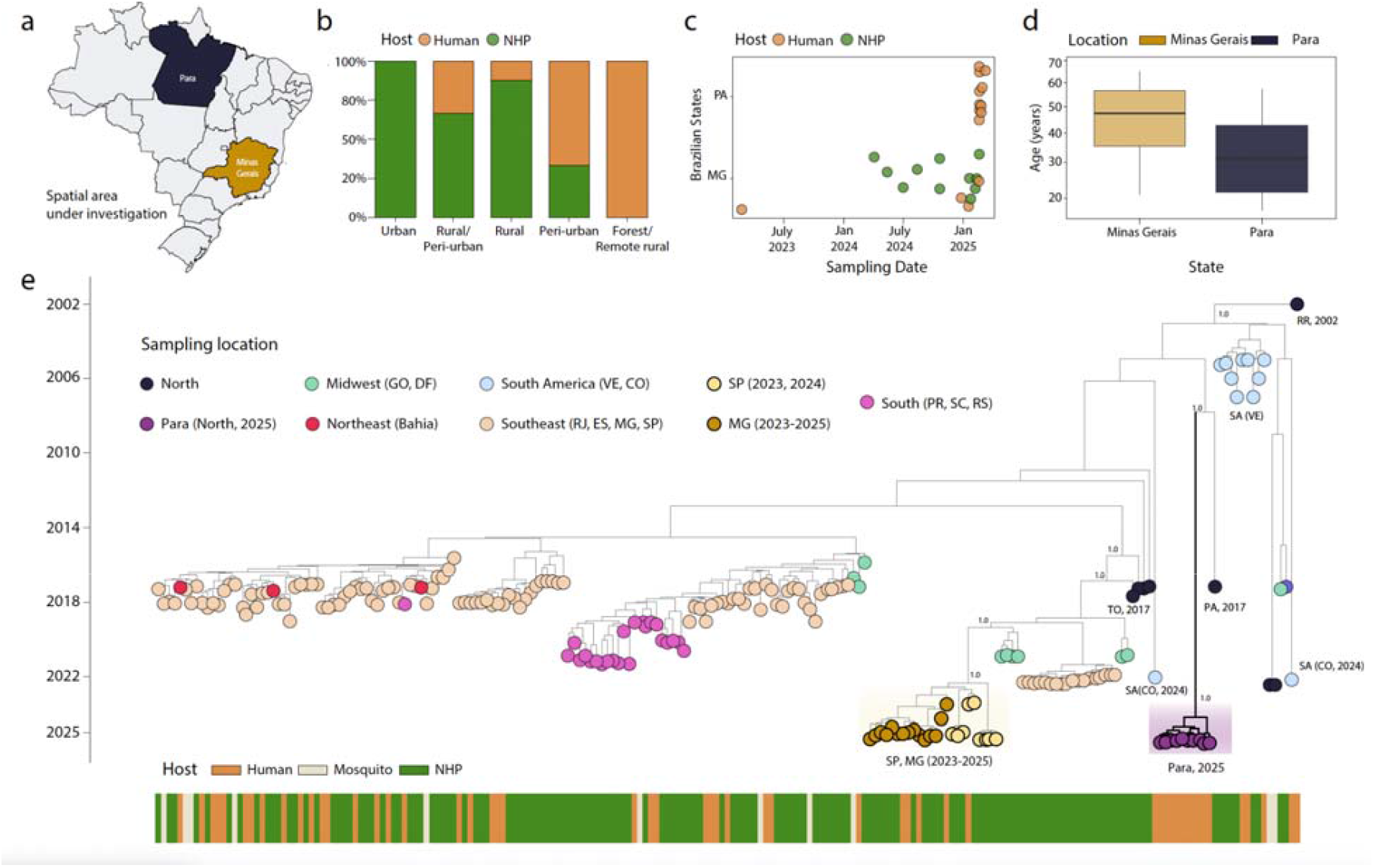
Genomic and epidemiological characterization of YFV cases in Pará and Minas Gerais, Brazil (2023–2025). a) Map of Brazil indicating the states of Pará (dark blue) and Minas Gerais (gold) under investigation; b) Proportion of YFV genomes by host (human: orange; NHP: green) and ecological classification (urban, rural/peri-urban, rural, peri-urban, forest/remote rural); c) Sampling dates of sequenced genomes by state (PA and MG) and host; d) Age distribution of human cases by state.; e) Time-scaled maximum likelihood phylogeny including genomes generated in this study and publicly available sequences. Tip colors indicate sampling location; tip fill denotes host (human: orange; NHP: green; mosquito: beige). Vertical bars to the right summarize host metadata. All sequences belong to the South American I lineage.

Ecological classification of the sampling sites indicated that all human infections identified in Pará were associated with forested or remote rural environments. In contrast, viral genomes recovered from Minas Gerais were predominantly obtained from NHPs across a broader range of ecological contexts, including rural, peri-urban, and urban areas. These findings likely reflect regional differences in host surveillance and sampling coverage rather than true ecological distribution. Notably, the exclusive detection of human infections in Pará is of epidemiological significance given the limited documentation of YFV human cases in this northern region in recent years (**Figure 2b, Table 1**). This is further supported by active surveillance near the municipality of Breves, where health authorities reported NHP carcasses and accounts from local residents of primate mortality occurring 2–3 months before human cases. Such observations underscore the role of NHP epizootic surveillance as a critical early warning tool for YFV circulation (Brazilian Ministry of Health, 2025). However, implementing this strategy in the Amazon remains challenging due to dense vegetation and limited access, which can hinder timely sample collection and transport.

Temporal analysis of the newly generated genomes indicated sustained YFV circulation in Minas Gerais from early 2023 to early 2025, whereas in Pará YFV genome detections were restricted to a shorter period, with all sequences clustering in 2025 (**Figure 2c**). Demographic data from confirmed human infections (n = 13) showed that all cases occurred in males aged between 17 and 65 years. Comparative analysis revealed a higher median age among infected individuals in Minas Gerais (∼52 years) relative to those in Pará (∼39 years), although this difference was not statistically significant (t-test p = 0.32, **Figure 2d**) likely due to the small number of data points available for Minas Gerais (n=4). Sequencing quality varied across samples, with Ct values ranging from 11 to 35. Genome coverage exceeded 85% in 19 of the 29 sequences, and as expected, depth of coverage showed an inverse correlation with Ct values (**Table 1**).

All genomes generated in this study were classified within the South American I lineage (**Figure 2e**). Sequences from Pará formed a well-supported monophyletic group, with Bayesian phylogenetic analysis estimating its emergence around mid-March 2024 (95% highest posterior density [HPD]: early March 2023 to October 2024). Notably, this clade was rooted by a basal strain from Pará collected in 2017, suggesting possible undetected viral persistence and subsequent detected re-emergence in the region. In contrast, sequences from Minas Gerais clustered with recent genomes from São Paulo (Cunha et al., 2025), forming a distinct lineage closely related to a genome from Goiás (Midwest Brazil). The estimated time to the most recent common ancestor (tMRCA) for this clade was late February 2019 (HPD: mid-January 2018 to mid-July 2019). This lineage was genetically distinct from strains associated with the 2016–2018 southeastern outbreak, supporting a scenario of a more recent reintroduction likely around early 2022, followed by sustained local transmission and diversification (Cunha et al., 2025).

Together, these findings provide genomic and epidemiological evidence for the co-circulation of distinct YFV lineages in Brazil during a period of expansion in viral activity, highlighting patterns of persistence and re-emergence in the North of the country, alongside sustained endemic transmission in the Southeast. Integrated genomic surveillance remains critical to elucidate region-specific transmission dynamics and inform timely, geographically tailored public health responses to mitigate the risk of future outbreaks. The findings of this report further underscore the currently existing risk of YFV re-establishing an endemic transmission cycle, supported by both a high abundance of Aedes aegypti and outdated 17DD vaccination coverages across the country.

## Data Availability

NA

## Author Contributions

Conceptualization: MG and LCJA; Methodology: VGDDA; TERA; VF; KMFM; LMRT; LAP; DGLDLR; AMBDF; DGR; DBR; KCLF; GALB; LCM; LCVF; LOL; NRG; PMSSB; MSDO; PSDA; PESDS; RGP; RGS; SMDSC; SHSPP; SK; SGP; VLDS; JL; FCDMI; ASJJ; MG; LCJA; Investigation: VGDDA; TERA; VF; KMFM; LMRT; LAP; DGLDLR; AMBDF; DGR; DBR; KCLF; GALB; LCM; MSDO; LCVF; LOL; NRG; PMSSB; PSDA; PESDS; RGP; RGS; SMDSC; SHSPP; SK; SGP; VLDS; JL; FCDMI; ASJJ; MG; LCJA; Data curation: VF, LMRT, and MG; Original draft preparation: TERA, VF, JL and MG; Visualization: VF; JL and MG. All authors have read and agreed to the published version of the manuscript.

## Funding Declaration

This study was supported by the National Institutes of Health USA grant U01 AI151698 for the United World Arbovirus Research Network (UWARN) and by the Novo Nordisk Foundation (NNF24OC0094346).

## Conflict of Interest

None declared.

## Notes

### Competing Interest Statement

The authors have declared no competing interest.

### Author Declarations

This project was reviewed and approved by the Pan American Health Organization Ethics Review Committee (PAHOERC) (Ref. No. PAHO 2016 08 0029) and the Oswaldo Cruz Foundation Ethics Committee (CAAE: 90249218.6.1001.5248). The availability of these samples for research purposes during outbreaks of national concern is allowed to the terms of the 510 2016 Resolution of the National Ethical Committee for Research Brazilian Ministry of Health (CONEP Comissao Nacional de Etica em Pesquisa, Ministerio da Saude), that authorize, without the necessity of an informed consent, the use of clinical samples collected in the Brazilian Central Public Health Laboratories to accelerate knowledge building and contribute to surveillance and outbreak response. The samples processed in this study were obtained anonymously from material exceeding the routine diagnosis of arboviruses in Brazilian public health laboratories that belong to the public network within BrMoH.

